# FTLD targets brain regions expressing recently evolved genes

**DOI:** 10.1101/2023.10.27.23297687

**Authors:** Lorenzo Pasquini, Felipe L. Pereira, Sahba Seddighi, Yi Zeng, Yongbin Wei, Ignacio Illán-Gala, Sarat C. Vatsavayai, Adit Friedberg, Alex J. Lee, Jesse A. Brown, Salvatore Spina, Lea T. Grinberg, Daniel W. Sirkis, Luke W. Bonham, Jennifer S. Yokoyama, Adam L. Boxer, Joel H. Kramer, Howard J. Rosen, Jack Humphrey, Aaron D. Gitler, Bruce L. Miller, Katherine S. Pollard, Michael E. Ward, William W. Seeley

**Affiliations:** Department of Neurology, Memory and Aging Center, University of California, San Francisco, CA, USA; Department of Neurology, Neuroscape, University of California, San Francisco, CA, USA; National Institute of Neurological Disorders and Stroke, Bethesda, MD, USA; Department of Genetics, Stanford University School of Medicine, Stanford, CA, USA; School of Artificial Intelligence, Beijing University of Posts and Telecommunications, Beijing, China; Global Brain Health Institute, University of California, San Francisco, San Francisco, CA, USA and Trinity College Dublin, Dublin, Ireland; Department of Neurology, Hospital de la Santa Creu i Sant Pau, Biomedical Research Institute, Universitat Autònoma de Barcelona, Barcelona, Catalunya, Spain; Department of Pathology, University of California, San Francisco, CA, USA; Department of Radiology, University of California, San Francisco, CA, USA; Nash Family Department of Neuroscience and Friedman Brain Institute, Icahn School of Medicine at Mount Sinai, New York, NY, USA; Gladstone Institute of Data Science and Biotechnology, San Francisco, CA, USA; Institute for Human Genetics, University of California San Francisco, San Francisco, CA, USA; Department of Epidemiology & Biostatistics and Bakar Institute for Computational Health Sciences, University of California San Francisco, San Francisco, CA, USA; Chan Zuckerberg Biohub, San Francisco, CA, USA

**Keywords:** Frontotemporal lobar degeneration, cryptic exon, human accelerated regions, TDP-43, tau, gene expression

## Abstract

In frontotemporal lobar degeneration (FTLD), pathological protein aggregation is associated with a decline in human-specialized social-emotional and language functions. Most disease protein aggregates contain either TDP-43 (FTLD-TDP) or tau (FTLD-tau). Here, we explored whether FTLD targets brain regions that express genes containing human accelerated regions (HARs), conserved sequences that have undergone positive selection during recent human evolution. To this end, we used structural neuroimaging from patients with FTLD and normative human regional transcriptomic data to identify genes expressed in FTLD-targeted brain regions. We then integrated primate comparative genomic data to test our hypothesis that FTLD targets brain regions expressing recently evolved genes. In addition, we asked whether genes expressed in FTLD-targeted brain regions are enriched for genes that undergo cryptic splicing when TDP-43 function is impaired. We found that FTLD-TDP and FTLD-tau subtypes target brain regions that express overlapping and distinct genes, including many linked to neuromodulatory functions. Genes whose normative brain regional expression pattern correlated with FTLD cortical atrophy were strongly associated with HARs. Atrophy-correlated genes in FTLD-TDP showed greater overlap with TDP-43 cryptic splicing genes compared with atrophy-correlated genes in FTLD-tau. Cryptic splicing genes were enriched for HAR genes, and vice versa, but this effect was due to the confounding influence of gene length. Analyses performed at the individual-patient level revealed that the expression of HAR genes and cryptically spliced genes within putative regions of disease onset differed across FTLD-TDP subtypes. Overall, our findings suggest that FTLD targets brain regions that have undergone recent evolutionary specialization and provide intriguing potential leads regarding the transcriptomic basis for selective vulnerability in distinct FTLD molecular-anatomical subtypes.

## Introduction

Frontotemporal dementia (FTD) refers to a group of clinical syndromes linked to underlying frontotemporal lobar degeneration (FTLD) pathology. These FTD syndromes present with deficits in recently evolved social-emotional and language functions^1,2^, resulting from degeneration within networks anchored by frontal, insular, and anterior temporal brain regions^3,4^. The most common FTD syndromes are the behavioral variant (bvFTD) characterized by social-emotional deficits^2^; semantic variant primary progressive aphasia (svPPA), which presents with loss of knowledge related to words, objects, and emotions^1,5^; nonfluent variant PPA (nfvPPA), characterized by speech production difficulties^1,5^; corticobasal syndrome, an asymmetric akinetic-rigid syndrome associated with a progressive loss of limb controls^4^; and progressive supranuclear palsy-Richardson syndrome (PSP-RS), associated with oculomotor deficits and gait instability^4^. FTLD pathology, in turn, can be divided into three major molecular classes based whether the neuronal and glial inclusions are composed of tau (FTLD-tau), TDP-43 (FTLD-TDP) or FUS/EWS/TAF15 (FET) family proteins^6^. Each major molecular class can be further divided into distinct subtypes based on the pathomorphology and distribution of the inclusions across regions, layers, and cell types. Factors driving the selective predilection of each FTLD subtype for distinct, yet overlapping, anatomical structures remain unknown.

Comparative genomic studies have the potential to shed light on the relationship between brain evolution and the erosion of human-specialized functions in FTLD. Novel genome-wide studies have identified a unique set of loci that are conserved across mammalian evolution, indicating important biological roles, but significantly diverged in the human lineage, suggesting changes to those roles in humans compared to chimpanzees and other primates^7,8^. These short sequences, referred to as human accelerated regions (HARs), are commonly located in non-coding DNA, often near genes associated with transcription and DNA binding^9^. Several studies have linked HARs to the emergence of distinctive human traits such as the opposable thumb^8^, language, and social behavior^10^, and mutations in HAR genes are observed in neuropsychiatric^11,12^ and neurodevelopmental disorders^7^.

Because FTLD is pathologically heterogeneous, it is possible that FTLD-TDP and FTLD-tau intersect with brain evolution through distinct yet overlapping mechanisms. The role of protein aggregation has been a major focus of research, but in FTLD-TDP, TDP-43 aggregation is almost always accompanied, and may be preceded, by loss of nuclear TDP-43 expression^13–16^. TDP-43, encoded by *TARDBP*, is an RNA-binding protein predominantly expressed in the nucleus of healthy neurons^17–19^, where it plays diverse roles in transcription regulation. One role is to suppress incorporation of cryptic exons, short stretches of non-conserved intronic RNA that are normally removed during pre-mRNA splicing^20–23^. Upon TDP-43 loss-of-function, cryptic exons are spliced into messenger RNAs, often introducing premature stop codons or frame shifts that result in nonsense mediated decay and loss of normal protein expression^24–26^. To date, more than 1,000 potential genes subject to cryptic splicing (CS) have been identified, yet, despite some compelling candidates^17,24^, it remains unknown which CS events, if any, are most relevant to FTLD-TDP pathogenesis. Crucially, although TDP-43 is a highly conserved protein, CS genes show almost no overlap when comparing mice to humans^20^, suggesting that TDP-43 may have adopted a unique regulatory role across the human evolutionary lineage.

Here, we combined neuroimaging, neuropathological, normative regional brain transcriptomic, comparative genomic, and cell biological data to identify genes whose expression pattern, in the healthy brain, correlates with cortical atrophy patterns in FTLD and to test the hypothesis that HAR genes are expressed in patterns resembling FTLD atrophy. We further explored associations between expression patterns for TDP-43 CS genes and FTLD-associated atrophy and the overlap between HAR and CS genes.

## Methods

### Participants

We searched the University of California, San Francisco (UCSF) Neurodegenerative Disease Brain Bank database for participants autopsied between 2008 and 2020 who had at least one high quality structural MRI scan prior to death; this search yielded 322 participants. From this pool, we included patients with a primary neuropathological diagnosis of FTLD-TDP (Types A-C) or FTLD-tau (corticobasal degeneration [CBD] or Pick’s disease). Although PSP is a common subtype of FTLD-tau, we omitted it here because it is characterized by a predominantly subcortical atrophy pattern not well suited to our transcriptomic data (see below). Patients were excluded if they had: (*i*) Braak neurofibrillary tangle stage ^27^ □>□3, (*ii*) Alzheimer’s disease neuropathological change ^28^ □>□intermediate, (*iii*) Lewy body disease ^29^ □>□brainstem predominant, or (*iv*) major territorial ischemic infarcts or intracranial hemorrhages^30^. Based on these stringent criteria, 28 patients with TDP-A, 35 with TDP-B, 29 with TDP-C, 45 with CBD, and 27 with Pick’s disease were included in the study. In accordance with the declaration of Helsinki, patients or their surrogates provided written informed consent prior to participation, including consent for brain donation. The UCSF Committee on Human Research approved the study.

All patients received clinical diagnoses by a multidisciplinary team following thorough neurological, neuroimaging, and neuropsychological assessments. Clinical severity was assessed using the Mini Mental State Examination^31^ and the Clinical Dementia Rating scale total and sum of boxes scores, using a version of the Clinical Dementia Rating scale adapted for FTD^32^. Patient demographics, as well as clinical and neuropathological data, are provided in **Table 1**. Race and ethnicity, handedness, and years of education were self-reported by patients or surrogates.

**Table 1.**
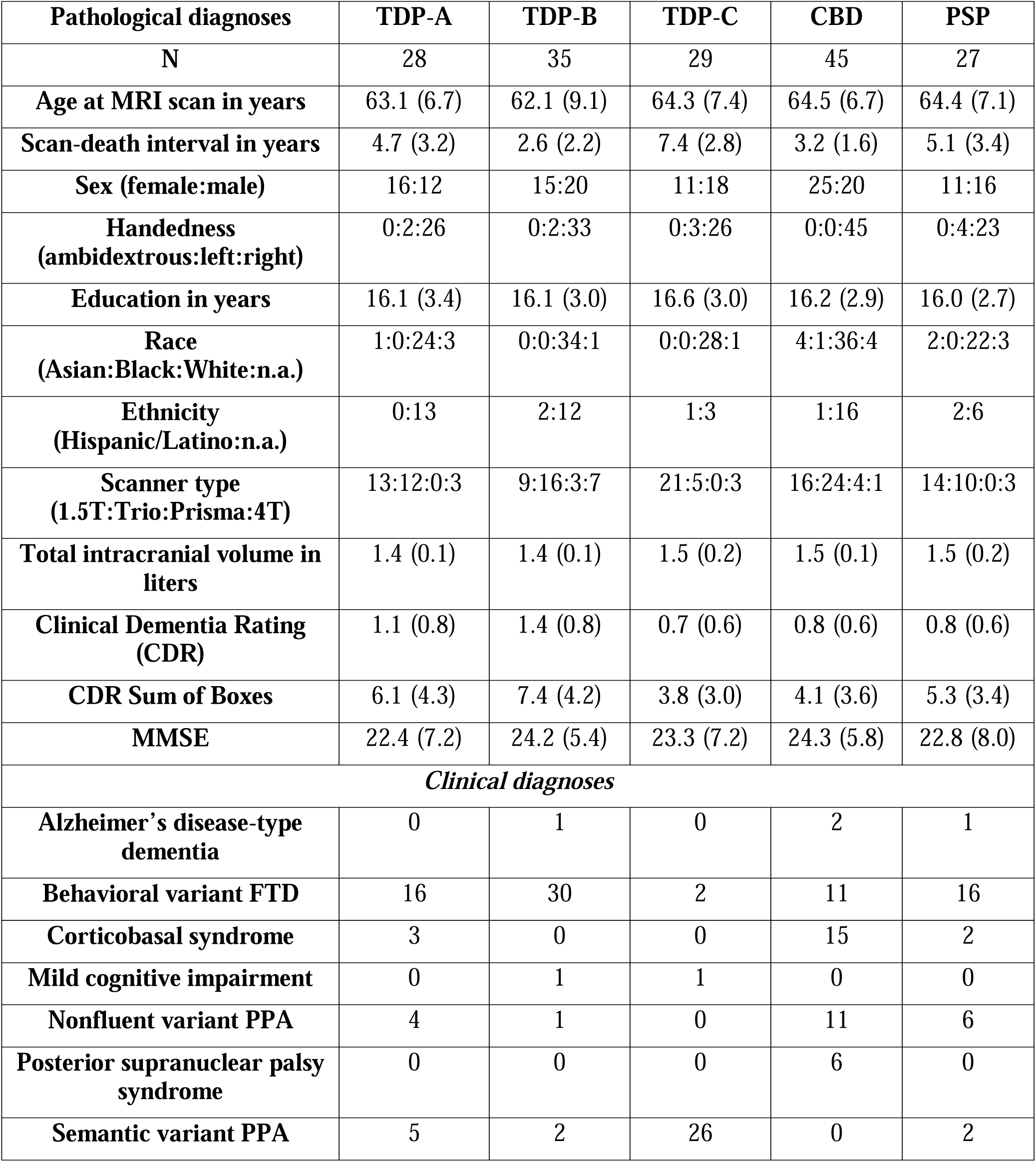
Pathological, clinical, and demographical characteristics of patients with FTLD. FTD = frontotemporal dementia; MMSE = Mini Mental State Examination; n.a. = not available; PPA = primary progressive aphasia

| **Pathological diagnoses** | **TDP-A** | **TDP-B** | **TDP-C** | **CBD** | **PSP** |
| --- | --- | --- | --- | --- | --- |
| **N** | 28 | 35 | 29 | 45 | 27 |
| **Age at MRI scan in years** | 63.1 (6.7) | 62.1 (9.1) | 64.3 (7.4) | 64.5 (6.7) | 64.4 (7.1) |
| **Scan-death interval in years** | 4.7 (3.2) | 2.6 (2.2) | 7.4 (2.8) | 3.2 (1.6) | 5.1 (3.4) |
| **Sex (female:male)** | 16:12 | 15:20 | 11:18 | 25:20 | 11:16 |
| **Handedness**<br>**(ambidextrous:left:right)** | 0:2:26 | 0:2:33 | 0:3:26 | 0:0:45 | 0:4:23 |
| **Education in years** | 16.1 (3.4) | 16.1 (3.0) | 16.6 (3.0) | 16.2 (2.9) | 16.0 (2.7) |
| **Race**<br>**(Asian:Black:White:n.a.)** | 1:0:24:3 | 0:0:34:1 | 0:0:28:1 | 4:1:36:4 | 2:0:22:3 |
| **Ethnicity**<br>**(Hispanic/Latino:n.a.)** | 0:13 | 2:12 | 1:3 | 1:16 | 2:6 |
| **Scanner type**<br>**(1.5T:Trio:Prisma:4T)** | 13:12:0:3 | 9:16:3:7 | 21:5:0:3 | 16:24:4:1 | 14:10:0:3 |
| **Total intracranial volume in**<br>**liters** | 1.4 (0.1) | 1.4 (0.1) | 1.5 (0.2) | 1.5 (0.1) | 1.5 (0.2) |
| **Clinical Dementia Rating**<br>**(CDR)** | 1.1 (0.8) | 1.4 (0.8) | 0.7 (0.6) | 0.8 (0.6) | 0.8 (0.6) |
| **CDR Sum of Boxes** | 6.1 (4.3) | 7.4 (4.2) | 3.8 (3.0) | 4.1 (3.6) | 5.3 (3.4) |
| **MMSE** | 22.4 (7.2) | 24.2 (5.4) | 23.3 (7.2) | 24.3 (5.8) | 22.8 (8.0) |
| ***Clinical diagnoses*** |  |  |  |  |  |
| **Alzheimer's disease-type**<br>**dementia** | 0 | 1 | 0 | 2 | 1 |
| **Behavioral variant FTD** | 16 | 30 | 2 | 11 | 16 |
| **Corticobasal syndrome** | 3 | 0 | 0 | 15 | 2 |
| **Mild cognitive impairment** | 0 | 1 | 1 | 0 | 0 |
| **Nonfluent variant PPA** | 4 | 1 | 0 | 11 | 6 |
| **Posterior supranuclear palsy**<br>**syndrome** | 0 | 0 | 0 | 6 | 0 |
| **Semantic variant PPA** | 5 | 2 | 26 | 0 | 2 |

### Neuropathology

*Postmortem* research-oriented autopsies were performed at the UCSF Neurodegenerative Disease Brain Bank. Neuropathological diagnoses were made following consensus diagnostic criteria^6,29,33^ based on standard histological and immunohistochemical methods^34,35^.

### Structural Neuroimaging

#### Acquisition and preprocessing

Because patients were evaluated over an 18-year interval, MRI scans were obtained on different scanners over time. For 74 patients, T1-weighted magnetization prepared rapid gradient echo (MPRAGE) MRI sequences were acquired at the UCSF Neuroscience Imaging Center, either on a 3T Siemens Tim Trio (n=67) or a 3T Siemens Prisma Fit scanner (n=7). Both scanners had similar acquisition parameters (sagittal slice orientation; slice thickness = 1.0 mm; slices per slab = 160; in-plane resolution = 1.0×1.0 mm; matrix = 240×256; repetition time = 2,300 ms; inversion time = 900 ms; flip angle = 9°), although echo time slightly differed (Trio: 2.98 ms; Prisma: 2.9 ms). For the remaining 87 patients, MRI scans were obtained at the San Francisco Veterans Affairs Medical Center using MPRAGE sequences acquired either on a Siemens 1.5 Tesla Magnetom scanner (n=73; voxel resolution 1.0 × 1.0 × 1.5 mm; TR□=□10 ms; TE□=□4 ms; TI□=□300 ms; flip angle□=□15) or on a Siemens 4 Tesla Magnetom scanner (n=14; voxel resolution 1.0 × 1.0 × 1.0 mm; TR□=□2.3 ms; TE□=□3 ms; TI□=□950 ms; flip angle□=□7). The average interval between scanning date and autopsy was 4.8 +/- 2.6 years.

All structural MR images underwent a voxel-based morphometry analysis^36^ after being visually inspected for motion and scanning artifacts. Structural images were segmented in gray matter, white matter, and cerebrospinal fluid and normalized to MNI space using SPM12 (http://www.fil.ion.ucl.ac.uk/spm/software/spm12/). Gray matter images were modulated by dividing the tissue probability values by the Jacobian of the warp field. The resulting images of voxel resolution 2.0 x 2.0 x 2.0 mm were smoothed with an isotropic Gaussian kernel with a full width at half maximum of 8 mm.

#### W-score maps

To generate participant-specific atrophy maps, the smoothed gray matter images were transformed to W-score maps. W-score maps are voxel-wise statistical maps that reflect levels of atrophy for each individual after adjustment for relevant covariates^37,38^. This approach is particularly useful when performing analyses with heterogenous samples since gray matter atrophy estimates are adjusted for demographical and methodological nuisance covariates, including age, gender, scanner type, and total intracranial volume^38^. The W-score model used for this study was published^39^; it involved 397 healthy older controls and included age at MRI, sex, years of education, handedness, scanner type, and total intracranial volume as covariates. Briefly, multiple regression analyses at the voxel-wise level were performed on the normative dataset to estimate gray matter intensity as adjusted for age, sex, years of education, handedness, MRI scanner, and total intracranial volume. This model was then used to estimate the expected gray matter intensity for a patient by using their demographical data. The discrepancy between the raw and the expected maps is used to derive the W-score map:

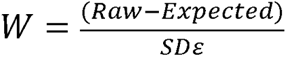

where *Raw* is the raw value of a voxel from the smoothed image of a patient; *Expected* is the expected value for the voxel of that specific patient based on the healthy control model; and *SD*ε is the standard deviation of the residuals from the healthy control model.

### Gene expression

#### Allen Human Brain Atlas (AHBA)

Normative human brain gene expression data were derived from microarray data available through the AHBA (http://human.brain-map.org/static/download). As described in detail elsewhere^40^, tissue samples were extracted across both hemispheres from two human brain donors, as well as the left hemisphere of four additional donors, totaling 3,702 tissue samples. Microarray analysis quantified gene expression across 58,692 probes, providing an estimate of the relative expression of 20,734 genes within the tissue samples^41^. The publicly available toolbox abagen (https://github.com/rmarkello/abagen)^42^ was then used for: *(i)* updating probe-to-gene annotations using the latest available data; *(ii)* data filtering, where expression values that do not exceed background are removed; *(iii)* probe selection, which, for genes indexed by multiple probes, involves selecting a single representative measure to represent the expression of that gene across all donor brains; *(iv)* sample assignment, where tissue samples from the AHBA were mapped to 273 parcels of the Brainnetome brain regional parcellation atlas (https://atlas.brainnetome.org/)^43^. This atlas was selected because it was derived using functional and structural anatomy and connectivity, and it includes paired homologous bilateral regions. Steps *i-iv* were followed by *(v)* normalization of expression measures to account for inter-individual differences and outlying values; *(vi)* gene-set filtering, to remove genes that are inconsistently expressed across the six brains; and *(vii)* averaging gene expression across donors. This procedure resulted in 15,655 genes (***Supplementary Data***) and their normative expression in 273 brain parcels. Finally, we removed subcortical regions (cerebellum, thalamus, basal ganglia, and hippocampus) due to substantial differences in their gene expression values when compared to cortical parcels^44^, yielding a 202 regions x 15,655 genes matrix (***Supplementary Figure S1***, ***Supplementary Data***).

#### HAR genes

HARs were taken from a comparative genomic analysis that identified, from a list of several recent publications, loci with accelerated divergence in humans when compared to chimpanzees^7^. A total of 2,737 HARs were identified, representing 2,164 unique HAR-associated genes^7^. Of these 2,164 HAR genes, 1,373 were identified as sufficiently expressed in the brain based on the AHBA brain-expressed gene dataset (probes used if they exceeded background signal for more than 50% of all samples) and used in our analyses, referred to simply as HAR genes^45^ (***Supplementary Data***).

#### CS genes

Three prior studies contributed to the list of TDP-43-dependent CS genes^24–26^. These studies were chosen because they identified CS targets either based on FTLD-TDP human *postmortem* tissue data^24^ or iPSC-derived neuronal cell lines^25,26^, omitting studies based on non-neuronal cells. For the tissue-based study, splicing analyses were performed on RNA-sequencing data from TDP-43-positive and TDP-43-negative neuronal nuclei isolated from frontal cortices of seven patients with FTLD-TDP. This approach identified 66 CS genes^24^, of which 63 genes exceeded the brain expression threshold imposed on the AHBA data. In a separate study, RNA sequencing was performed on human iPSC-derived cortical-like neurons, in which TDP-43 expression was reduced using Clustered Regularly Interspaced Short Palindromic Repeats interference (CRISPRi)^25^. Differential splicing and expression analyses based on this dataset followed by splice junction evaluation via visual inspection of sashimi plots revealed 107 genes with putative CS events, of which 97 also exceeded the brain expression threshold in the AHBA dataset. Finally, a third study performed a comprehensive analysis of differential splicing events in TDP-43 depleted versus control iPSC-derived neurons to develop a high-quality neuronal cryptic exon atlas^26^. This analysis revealed 233 CS genes, of which 201 exceeded the brain expression threshold in the AHBA brain-expressed gene dataset, as previously defined. By combining the two iPSC-derived neuronal CS gene sets, 216 unique CS genes were identified and combined with the *postmortem* tissue-derived CS genes to yield a final brain-expressed CS gene set of 257 unique genes (***Supplementary Data***).

#### Overlap of gene lists

We used Venn diagrams to visualize the overlap between distinct gene lists. To test whether gene lists overlapped to a degree higher than expected by chance, we used bootstrap-based tests. In this procedure, for example, a gene list of the same number of genes as the list of HAR genes is randomly drawn 5,000 times from the AHBA dataset of brain-expressed genes. For each of these randomly selected bootstrap gene lists, we then quantify the overlap with the CS gene list, resulting in a distribution of overlaps between CS genes and randomly selected gene lists. We next count how often the random overlap was equal to or higher than the observed overlap between the HAR and CS gene lists and derive a *p* value by calculating the proportion of all bootstrap gene lists with an overlap exceeding the observed overlap.

#### Spatial correlation analyses

We derived group-averaged, voxel-wise W-score maps for each FTLD pathological subtype (TDP-A, TDP-B, TDP-C, CBD, and Pick’s disease). For each subtype-averaged map, we then averaged the W-score values of voxels contained in each cortical Brainnetome parcel to derive a vector reflecting the level of regional cortical atrophy in each FTLD subtype. These FTLD subtype-specific atrophy vectors were then separately correlated with the cortical gene expression levels derived from the AHBA brain-expressed gene dataset using Pearson’s correlation coefficients. This procedure enabled us to assess the spatial similarity between each FTLD subtype atrophy pattern and the cortical expression levels of 15,655 brain-expressed genes from the AHBA^44,46,47^.

Because spatial autocorrelation inherent to neuroimaging data can inflate *p*-values in brain map analyses^48^, we corrected for this autocorrelation by applying a bootstrap-based approach (***Supplementary Figure S2***), in which 5,000 surrogate maps that preserve the autocorrelation properties of the FTLD atrophy maps were generated using the toolbox BrainSMASH (https://brainsmash.readthedocs.io/en/latest/approach.html). We then derived autocorrelation-corrected *p*-values by counting how often the correlation between surrogate atrophy maps and each gene map was equal to or higher than the true correlation and divided this number by the total number of bootstraps. Only False Discovery Rate (FDR) adjusted *p*-values < 0.05 were considered significant. This procedure enabled us to identify genes with spatial expression patterns resembling the FTLD atrophy maps.

To remove genes displaying spurious correlations to FTLD-TDP atrophy maps from subsequent analyses, we assessed how applying a threshold based on the absolute correlation value would affect the list of selected genes significantly correlating with FTLD-TDP atrophy maps. We applied absolute correlation thresholds between 0-0.5 in steps of 0.05 and derived an index assessing how each threshold impacted the ratio between the number of genes uniquely associated with each FTLD-TDP atrophy map and the number of genes that significantly correlated with at least two FTLD-TDP subtype atrophy maps:

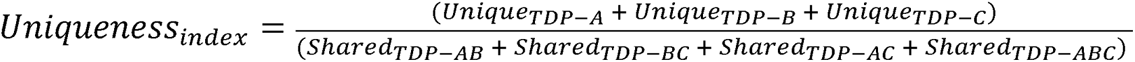

This uniqueness index was used to identify 0.2 as the optimal absolute correlation threshold to better identify subtype-specific correlating genes while at the same time preserving longer gene lists. Genes displaying correlation values between 0.2 and −0.2 were hence considered as not being sufficiently correlated with FTLD-TDP atrophy maps and discarded from further analyses regardless of the associated *p*-value. We then assessed the overlap between gene lists representing FTLD atrophy-correlated, HAR, and CS genes using the previously described bootstrap approach.

#### Gene set enrichment analysis

Gene set enrichment analyses are computational methods that query large genomic databases to determine whether an *a priori* defined set of genes shows statistically significant associations with known biological pathways. We ran five separate gene set enrichment analyses^49^ leveraging the correlation values (FDR adjusted *p* < 0.05) between brain genes and atrophy maps of FTLD-TDP and FTLD-tau subtypes (one analysis for each subtype). We performed gene set enrichment analyses using the *clusterProfiler* package in R software v4.2.1 for Gene Ontology^50^ and for Kyoto Encyclopedia of Genes and Genomes (KEGG)^51^. We then focused on terms and pathways with a Holm adjusted *p*-value of < 0.05 shared by a majority of FTLD subtypes. Correlation matrices were used to reflect the strength of correlation between genes associated with various KEGG terms and FTLD-TDP and FTLD-tau subtype atrophy patterns (FDR adjusted *p* < 0.05).

#### Graph network

Over the course of the study, we identified a set of genes that correlated with FTLD-TDP atrophy and overlapped with the previously introduced CS and HAR gene lists. Graph theoretical approaches were applied to this list of 23 CS-HAR genes correlating with FTLD-TDP atrophy to generate topographical representations of gene expression networks^52^. The regional expression values of selected genes were correlated with each other to generate a gene-to-gene regional co-expression matrix. This matrix was binarized at a Pearson’s correlation coefficient threshold of 0.3 and the number of surviving edges between genes was counted for each gene of interest to derive a measure of nodal degree, reflecting the level of connectedness between a specific gene and each other gene selected for the analysis. Resulting data were rendered as a graph.

#### Gene length

Standard *BiomatRt* code in R (https://useast.ensembl.org/info/data/biomart/index.html) leveraging the widely used Ensembl genome browser (https://ensemblgenomes.org/) was used to derive the full coordinates of exonic and intronic sequences of brain-expressed genes used in our analyzes (BSgenome.Hsapiens.UCSC.hg38). This information was used to estimate the length of HAR genes used in our analyses and to identify a set of 1,353 length-matched non-HAR genes through nearest neighbor matching using the *matchit* library in R.

#### Disease epicenters

We used an approach previously applied by our group and others^53–56^ to identify patient-tailored epicenters. This method defines an epicenter as the brain region whose normative functional (task-free) connectivity map most closely resembles the patient’s atrophy W-map. Normative connectivity maps were derived using task-free functional MRI data from a cohort of 75 healthy older subjects to generate a library of 194 intrinsic functional connectivity maps from seed regions of the Brainnetome atlas spanning the entire cerebral cortex. Characterization of the normative sample and methodological details are presented elsewhere^53^. We then compared each patient’s atrophy W-map with this library to select the seed that generated a functional connectivity map showing the highest spatial (Pearson) correlation to the subject-specific atrophy W-map. Subtype-specific frequency maps were used to quantify the spatial distribution of epicenters in FTLD-TDP, since group-level atrophy in FTLD-TDP was associated with both CS and HAR genes. Finally, we used Wilcoxon signed-rank tests to assess whether the normative expression of CS and HAR genes within disease epicenters differed across FTLD-TDP subtypes. For each gene, epicenters’ gene expression levels were then averaged for each subtype to derive a cross-fold change score reflecting higher or lower gene expression in one group versus the other.

## Results

### FTLD subtypes target brain regions expressing shared and distinct genes

We first sought to explore the relationship between FTLD atrophy topography and normative brain regional gene expression. We included the three major FTLD-TDP subtypes (A-C) and the two major sporadic FTLD-tau subtypes, CBD and Pick’s disease, with predominant cortical involvement. First, we sought to replicate the known atrophy patterns associated with these disorders using *in vivo* brain MRI data from patients with an autopsy-based diagnosis^57,58^. Structural MRI volumes were segmented into gray matter tissue probability maps and further transformed to W-score maps^37,38^, where higher W-scores reflect more severe voxel-wise atrophy for each patient, adjusted for demographic variables. FTLD subtype specific cortical atrophy maps were derived by averaging W-score maps within each group (**Figure 1A-E**, left panels), revealing a more dorsal frontal and insular pattern in TDP-A; a more ventral pattern in TDP-B, with less severe atrophy overall; an anterior temporal-predominant pattern in TDP-C; a milder, predominantly frontal atrophy pattern in CBD; and a severe frontal and insular pattern of atrophy in Picks’ disease.

**Figure 1.**
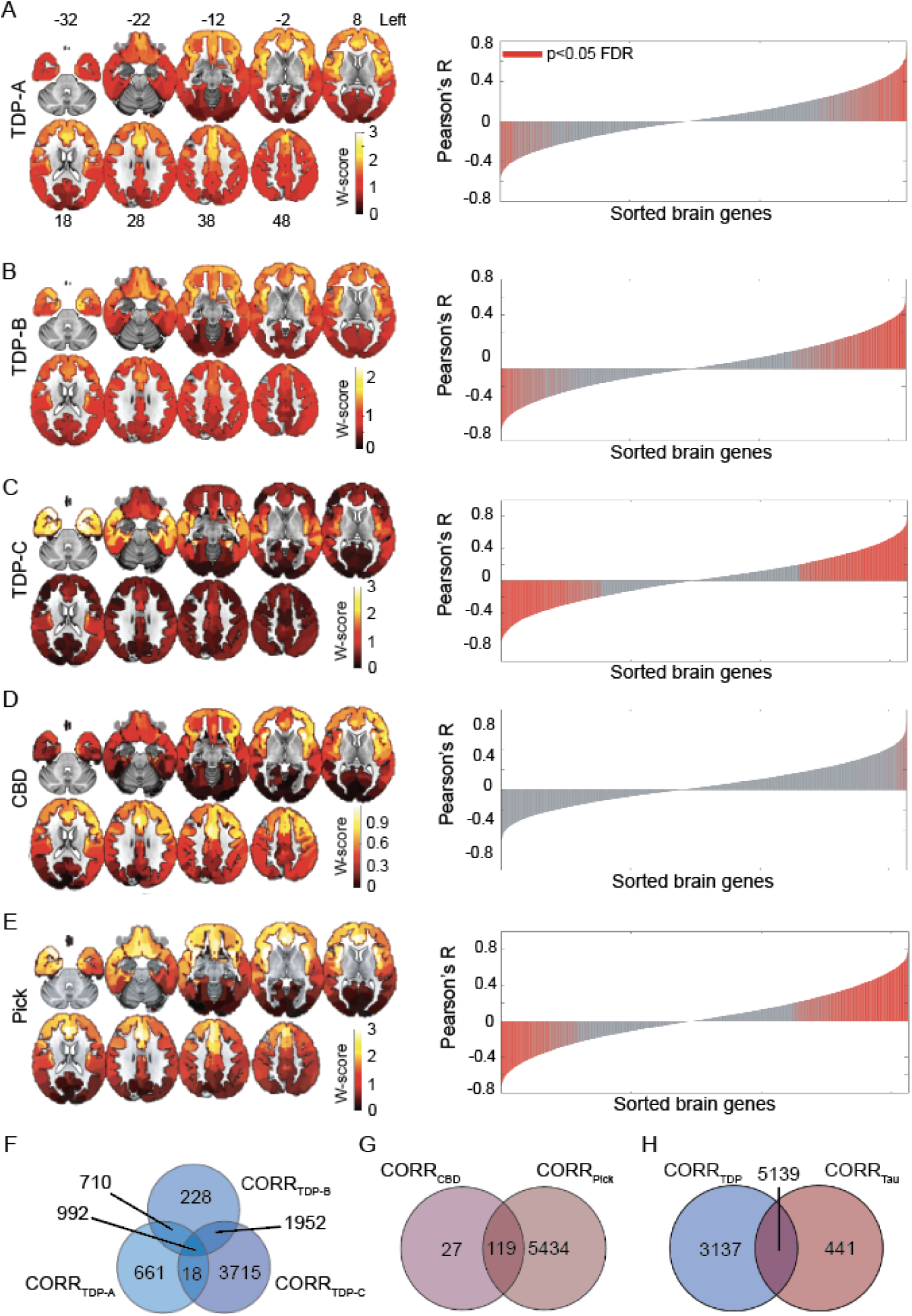
Regional gene expression correlating with atrophy in FTLD subtypes. **(A-E).** Maps on the left show group-averaged W-score maps for each FTLD subtype; warmer colors reflect greater cortical gray matter atrophy. Bar plots on the right show the Pearson correlation coefficients between the regional gene expression and regional atrophy in FTLD subtypes; red bars indicate significantly correlated genes, FDR adjusted *p* <0.05. **(F)** Several genes were shared among FTLD-TDP subtypes, although a conspicuous number of genes was uniquely correlated to each subtype, particularly to FTLD-TDP-C. **(G)** While FTLD-Pick showed many uniquely correlated genes, most genes correlating with atrophy in FTLD-CBD were shared with FTLD-Pick. **(H)** FTLD-TDP had many uniquely correlated genes, while a large proportion of genes correlating with atrophy in FTLD-tau were shared with FTLD-TDP. CORR_FTLD-subtype_ indicates lists of genes correlating with that specific FTLD subtype.

We next leveraged normative gene expression microarray data from the AHBA to identify genes whose cortical expression pattern resembles cortical gray matter atrophy characteristic of each FTLD subtype. Starting with a matrix of 15,655 genes and their regional expression in 202 cortical brain parcels (***Supplementary Figure S1***), we performed spatial correlation analyses to associate group-level gray matter atrophy patterns with normative regional expression levels of brain-expressed genes. We used a bootstrap-based test, correcting for the spatial autocorrelations inherent to neuroimaging data^48^, to delineate genes significantly correlated with atrophy in each FTLD subtype (FDR adjusted *p* < 0.05, **Figure 1A-E**, red bars in the right panels). To remove spurious correlations from further analyses, we only considered absolute correlation values ≥ 0.2, based on a uniqueness index maximizing the number of genes uniquely associated with each FTLD subtype (***Supplementary Figure S2***). This procedure identified unique and shared genes correlating with atrophy across FTLD subtypes. TDP-C showed the most uniquely correlated genes (3,715), followed by TDP-A (661 genes) and TDP-B (228 genes). A total of 992 genes were shared between all FTLD-TDP subtypes (**Figure 1F**). Among FTLD-tau, CBD showed only 27 uniquely correlated genes, due to the mild frontal atrophy found in this sample highly resembling the inherent frontal-posterior spatial autocorrelation gradient inherent to neuroimaging data^48^. CBD shared 119 genes with Pick’s disease, which yielded a total of 5,434 uniquely correlated genes for FTLD-tau (**Figure 1G**). When pooling both FTLD-TDP and FTLD-tau together, 3,137 genes were uniquely correlated with at least one TDP subtype, 441 with at least one tau subtype, and 5,139 were shared among TDP-43 and tau FTLD subtypes (**Figure 1H**).

To better understand the biological function of FTLD atrophy-correlated genes, we performed gene set enrichment analyses^49^ against gene ontology and Kyoto Encyclopedia of Genes and Genomes (KEGG)^50^. While no gene ontology was considered enriched, the gene set enrichment analysis revealed only one KEGG pathway, associated with neuroactive ligand-receptor interactions (has04080)^51^, shared across FTLD-TDP and FTLD-Pick subtypes. This KEGG pathway consisted of common and unique associations between FTLD atrophy-correlated genes and major classes of neuromodulatory and homeostatic neurotransmitter systems (**Figure 2**). Genes associated with acetylcholine receptors did not, for the most part, correlate with atrophy in any FTLD subtype (**Figure 2A**), as expected based on the lack of a cholinergic deficit in FTLD^59^. Conversely, genes associated with receptors within the epinephrine/norepinephrine, opioid, somatostatin, and neuropeptide Y systems correlated across most FTLD subtypes (**Figure 2B, F-H**). Some receptor systems showed more specific associations with FTLD subtypes, as in the case of serotonin and dopamine to TDP-C (**Figure 2C, D**), or oxytocin to TDP-B, TDP-C, and Pick’s disease (**Figure 2E)**. The lack of associations for CBD is likely attributable to a low number of genes associated with atrophy in this subtype and the high spatial autocorrelation of its atrophy map.

**Figure 2.**
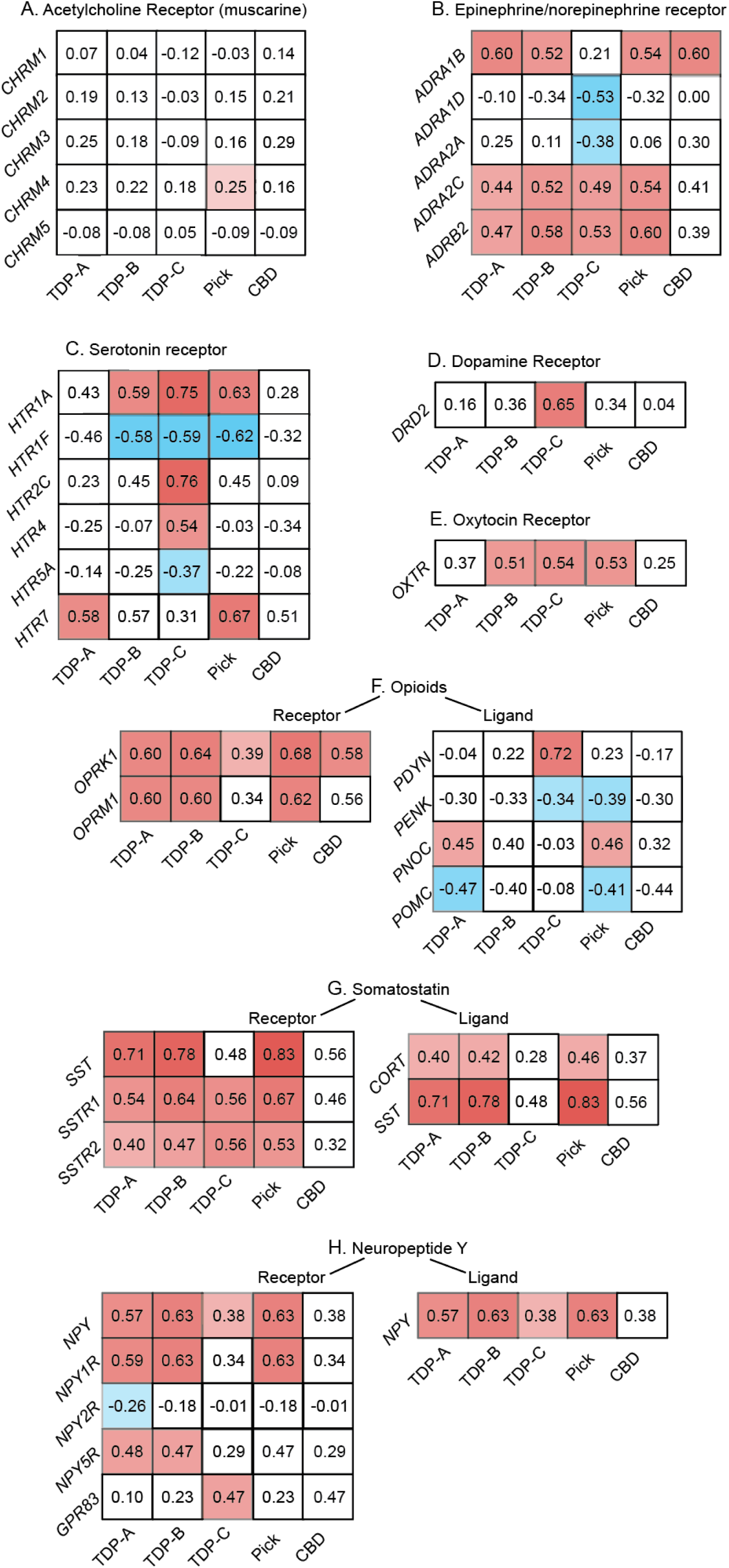
Genes correlating with atrophy in FTLD are enriched for neuromodulatory terms. Gene set enrichment analyses using the spatial correlation values between brain gene expression levels and FTLD atrophy severity revealed a KEGG neuroligand receptor interaction pathway in common among most FTLD subtypes. Whereas genes associated with acetylcholine neurotransmission **(A)**, a system often preserved in FTLD, did, for the most part, not correlate with FTLD atrophy, shared and unique associations were observed between FTLD subtypes and genes associated with epinephrine **(B)**, serotonin **(C)**, dopamine **(D)**, oxytocin **(E)**, opioids **(F)**, somatostatin **(G)**, and neuropeptide Y receptor and ligand systems **(H)**. Matrices reflect the strength and sign of spatial correlation between genes associated with ligand and receptor systems and atrophy maps in FTLD subtypes. White squares reflect genes whose correlation with FTLD atrophy maps did not reach significance based on the bootstrap test correcting for spatial autocorrelation (FDR adjusted *p* < 0.05).

### Genes correlated with FTLD atrophy are enriched for HAR genes

We identified 1,373 brain-expressed HAR genes derived from a comparative genomic study of conserved loci with elevated divergence in humans versus chimpanzees and other mammals (**Figure 3A, *Supplementary Materials***)^7^. We next assessed the overlap between brain-expressed HAR genes and genes whose expression patterns correlated with atrophy in FTLD. To determine whether these overlaps occurred at a rate greater than chance, we used a bootstrap-based test (***Supplementary Figure S3***), selecting random gene sets from the pool of brain-expressed genes. Atrophy-correlated genes were enriched for HAR genes in both FTLD-TDP (808 genes, *p* < 0.0005, **Figure 3B**) and FTLD-tau (560 genes, *p* < 0.0005, **Figure 3C**), with a significantly greater overlap observed for FTLD-TDP atrophy-correlated genes (*χ*^2^ = 89.59, *p* < 0.0001). HAR genes correlating with at least one FTLD subtype tended to be either unique to FTLD-TDP (283 genes) or shared between FTLD-TDP and FTLD-tau (525 genes) (***Supplementary Figure S4A***), with only 35 HAR genes correlating uniquely with at least one FTLD-tau subtype.

**Figure 3.**
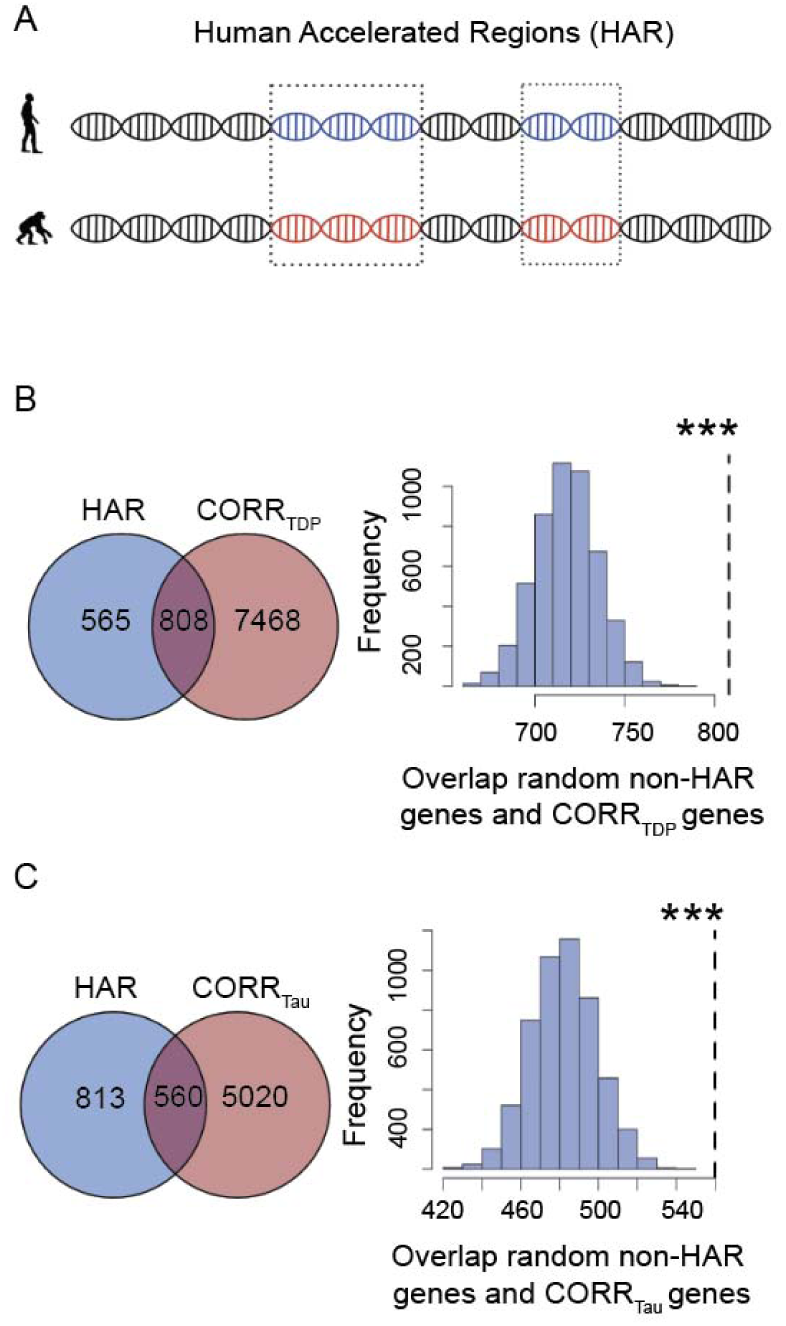
Overlap between FTLD atrophy correlated genes and HAR genes and CS genes. **(A)** HARs are conserved genomic loci which have undergone accelerated divergence in the human evolutionary lineage. **(B)** Genes correlating with atrophy in FTLD-TDP had 808 genes in common with HAR genes, **(C)** while genes correlating with atrophy in FTLD-tau had 560 genes in common with HAR genes. Both overlaps were unlikely to occur by chance when compared to a background set of brain-expressed genes. \*\*\**p* < 0.0005. Panel A adapted with permission from^7,45^.

### FTLD-TDP atrophy-correlated genes are linked to TDP-43 CS genes

Recent studies have identified a corpus of genes that undergo CS upon TDP-43 loss-of-function^24–26^ (**Figure 4A**). Given the strong enrichment of FTLD-TDP atrophy-correlated genes for HAR genes, and that humans show non-conserved patterns of TDP-43 related CS, we hypothesized that TDP-43 CS genes might be enriched among genes correlated with FTLD-TDP (but not FTLD-tau) atrophy. To explore this possibility, we consolidated published and unpublished TDP-43 CS gene lists, filtering for human brain-expressed CS genes identified using human cell models (216 genes)^25,26^ or patient brain tissues (63 genes^24^; ***Supplementary Table S1***), resulting in 257 unique CS genes (***Supplementary Materials***). 146 genes were shared between FTLD-TDP atrophy-correlated and CS genes, with this overlap trending towards significance (*p* = 0.10, **Figure 4B**). To evaluate the biological relevance of the observed trend, we compared the overlap between CS genes and FTLD-TDP atrophy-correlated genes to the overlap between CS genes and FTLD-tau atrophy-correlated genes. We identified 88 FTLD-tau correlated CS genes; in contrast to FTLD-TDP, however, this overlap did not approach significance (*p* = 0.71, **Figure 4C**). The proportion of CS genes overlapping with FTLD-TDP atrophy-correlated genes was significantly higher than the proportion of CS genes overlapping with FTLD-tau atrophy-correlated genes (*χ*^2^ = 26.39, *p* < 0.0001). To control these analyses for the smaller number of FTLD-tau atrophy-correlated genes, we randomly re-sampled a single list of 5,580 FTLD-TDP atrophy-correlated genes (matching the number of FTLD-tau atrophy-correlated genes), which revealed a significant overlap of 106 genes shared with CS genes (***Supplementary Figure S5****, p <* 0.05), suggesting that the differential overlap between FTLD-TDP and FTLD-tau atrophy-correlated genes with CS genes is not merely a reflection of gene list length. CS genes correlated with at least one FTLD subtype were either unique to FTLD-TDP (63 genes) or shared between FTLD-TDP and FTLD-tau (83 genes) (***Supplementary Figure S4B***). These finding suggest that although both FTLD-TDP and FTLD-tau target brain regions expressing recently evolved genes, FTLD-TDP exhibits a closer relationship to regions that express TDP-43 CS genes.

**Figure 4.**
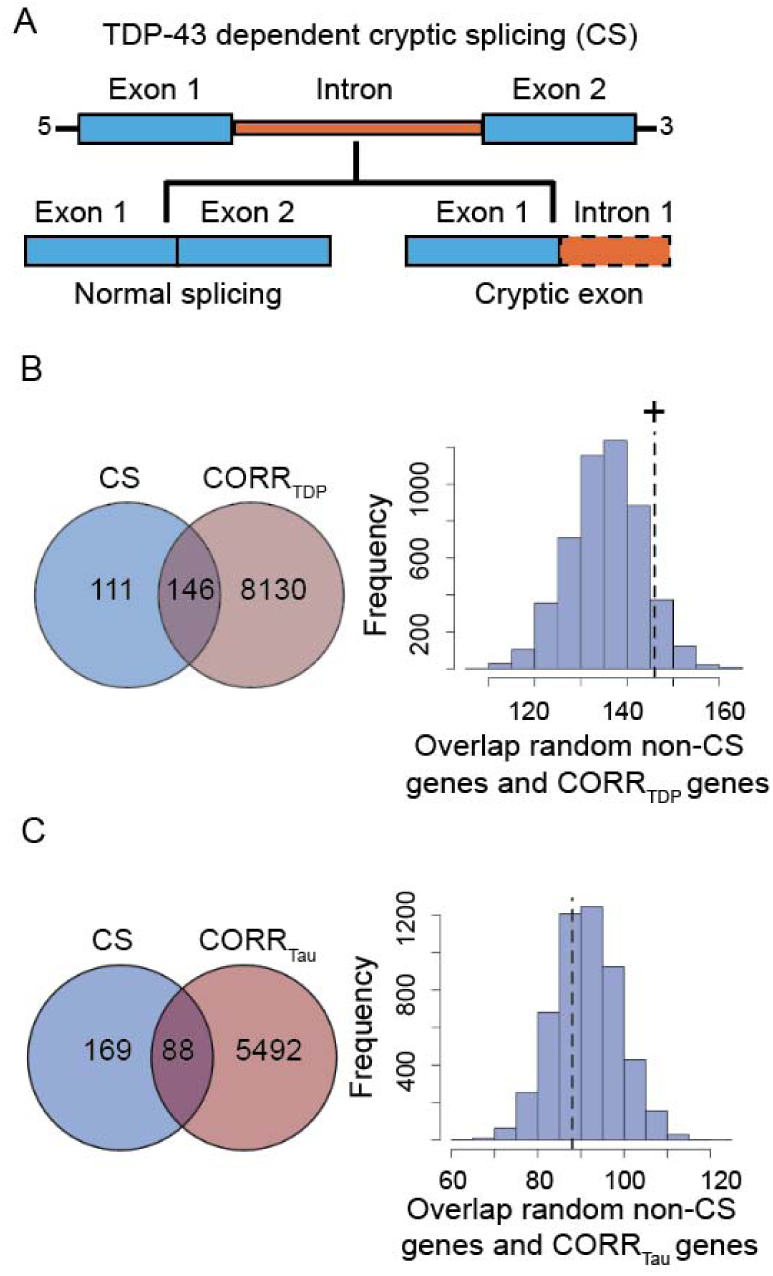
Overlap between FTLD atrophy correlated genes and HAR genes and CS genes. **(A)** CS genes are genes that incorporate novel intronic RNA into mature mRNA when TDP-43 is knocked down experimentally or depleted from diseased neuronal nuclei. **(B)** 146 genes were shared between CS genes and genes correlating with atrophy in FTLD-TDP. This overlap reached trend significance (*p* = 0.10). **(C)** 88 genes were shared between CS genes and genes correlating with atrophy in FTLD-tau, with this overlap being far from significant (*p* = 0.71). ^+^*p* < 0.1.

### HAR genes overlap with TDP-43 CS genes

Among the TDP-43 CS genes, 37 were also HAR genes (**Figure 5A, *Supplementary Materials***), an overlap that was higher than expected by chance, as demonstrated by distributions derived from bootstrap-based tests, where the overlap between randomly selected brain-expressed gene lists and HAR genes (*p* < 0.005) or CS genes (*p* < 0.0005) was repeatedly assessed and compared to the observed overlap between HAR genes and CS genes (**Figure 5B-C**). Identified CS-HAR genes included genes linked to social behavior and language (e.g., *ERC2*, *KIF26B*, *CBLN2, SEMA6D*)^60–64^, neurodevelopment, and neuropsychiatric diseases (e.g., *CBLN2SPRY3*, *CAMTA1*, *LRP8*)^65–69^. Some CS-HAR genes identified (e.g., *CBLN2*, *ICA1, PTPRT, and PTPRD*) have been shown to accurately distinguish FTLD-TDP *postmortem* cortex from non-neurological disease control tissue based on the frontal/temporal cortex expression of cryptic exons^26^. CS-HAR genes tended to be longer than other brain-expressed genes (*t*(39) = 4.74; *p* < 0.0001), but their length fell within the range found for 85% of brain expressed genes (***Supplementary Figure S6A***). CS-HAR genes contained mostly intronic HAR sequences (76%, ***Supplementary Figure S6B***), followed by intergenic (21%) and exonic (3%) sequences, and cryptic splicing events affecting CS-HAR genes mainly included cryptic exon incorporation^20^ (71%, ***Supplementary Figure S6C***), consistent with other (non-HAR) CS genes. We performed control analyses assessing the overlap between CS genes and a set of non-HAR genes matched for length to HAR genes (***Supplementary Figure S7A***). This approach yielded an overlap of 42 genes (***Supplementary Figure S7B***, χ^2^ = 0.36, *p* = 0.55), suggesting that gene size is a confounding factor driving the overlap between CS and HAR genes. Therefore, although HAR and TDP-43 CS genes overlap, our analyses do not provide evidence for a special, gene-length independent relationship between HAR and TDP-43 CS genes.

**Figure 5.**
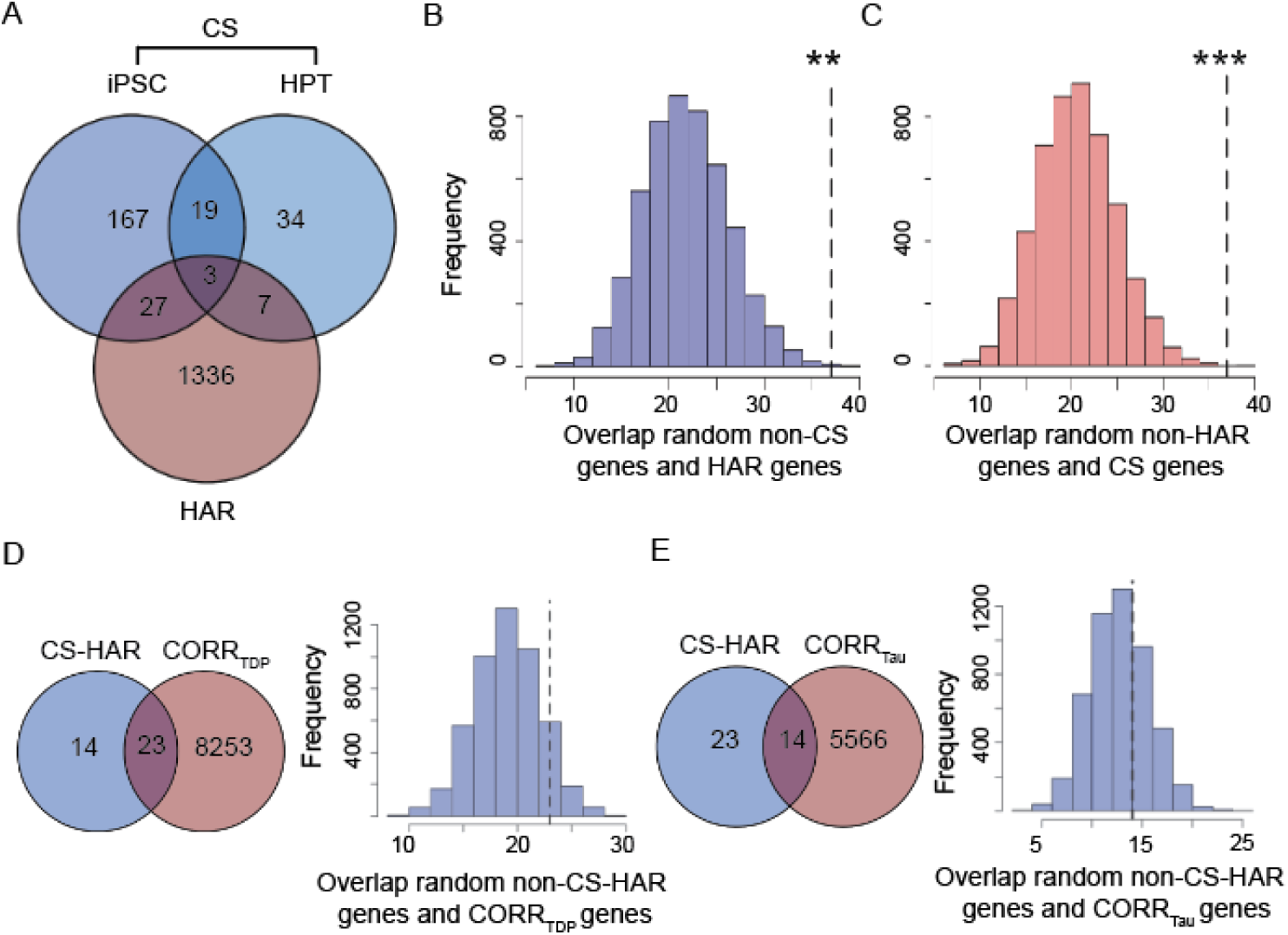
HAR and CS genes overlap. **(A)** Comparing the HAR and CS gene lists, including those derived from iPSC-derived neurons and from human patient tissues (HPT), 37 total genes were shared. Bootstrap-based testing, **(B-C)** was used to determine whether gene sets overlapped at a rate greater than expected by chance. First, we compared the overlap between random gene lists of the same number of genes as the CS genes with the HAR gene list **(B)**. Second, we compared the overlap between random gene lists of same number of genes as the HAR genes with the CS gene list **(C)**. Both tests showed that the overlap between CS genes and HAR genes is far greater than expected by chance. **(D)** 23 genes were shared between CS-HAR genes and genes correlating with atrophy in FTLD-TDP. This overlap was higher than expected by chance but did not reach nominal significance (*p* = 0.17). **(E)** 14 genes were shared between CS-HAR genes and genes correlating with atrophy in FTLD-tau, with this overlap not approaching significance (*p* = 0.46). \*\**p* < 0.005; \*\*\**p* < 0.0005.

We next sought to better understand relationships between FTLD atrophy-correlated genes and the 37 CS-HAR genes (**Figure 5A**). Of these 37 genes, 23 overlapped with FTLD-TDP atrophy-correlated genes (*p* = 0.17, **Figure 5D**) and 14 with FTLD-tau atrophy-correlated genes (*p* = 0.46, **Figure 5E**), but these overlaps were not greater than expected by chance. Nonetheless, the proportion of CS-HAR genes overlapping with FTLD-TDP atrophy-correlated genes was significantly higher than the proportion of CS-HAR genes overlapping with FTLD-tau atrophy-correlated genes (*χ*^2^ = 4.38, *p* < 0.05). CS-HAR genes correlating with at least one FTLD subtype tended to be either unique to FTLD-TDP (10 genes) or shared between FTLD-TDP and FTLD-tau (13 genes) (***Supplementary Figure S4C***). Accordingly, to generate hypotheses, we examined the relationships among the 23 FTLD-TDP atrophy-associated CS-HAR genes in terms of their co-expression relationships in the healthy brain and their links to specific FTLD-TDP subtypes (**Figure 6**). While some genes correlated negatively with atrophy (e.g., *PTPRD*), suggesting lower normative expression among FTLD-TDP atrophied regions, other genes correlated positively (e.g., *ERC2*), reflecting higher expression among targeted regions. Some FTLD-TDP subtype-specific associations emerged, such as *PTPRD* with TDP-A, *RHOBTB3* with TDP-B, or *ERC2* with TDP-C, while other genes, such as *RALGAPA2*, *ICA1*, and *SHISA9,* were shared among all FTLD-TDP subtypes.

**Figure 6.**
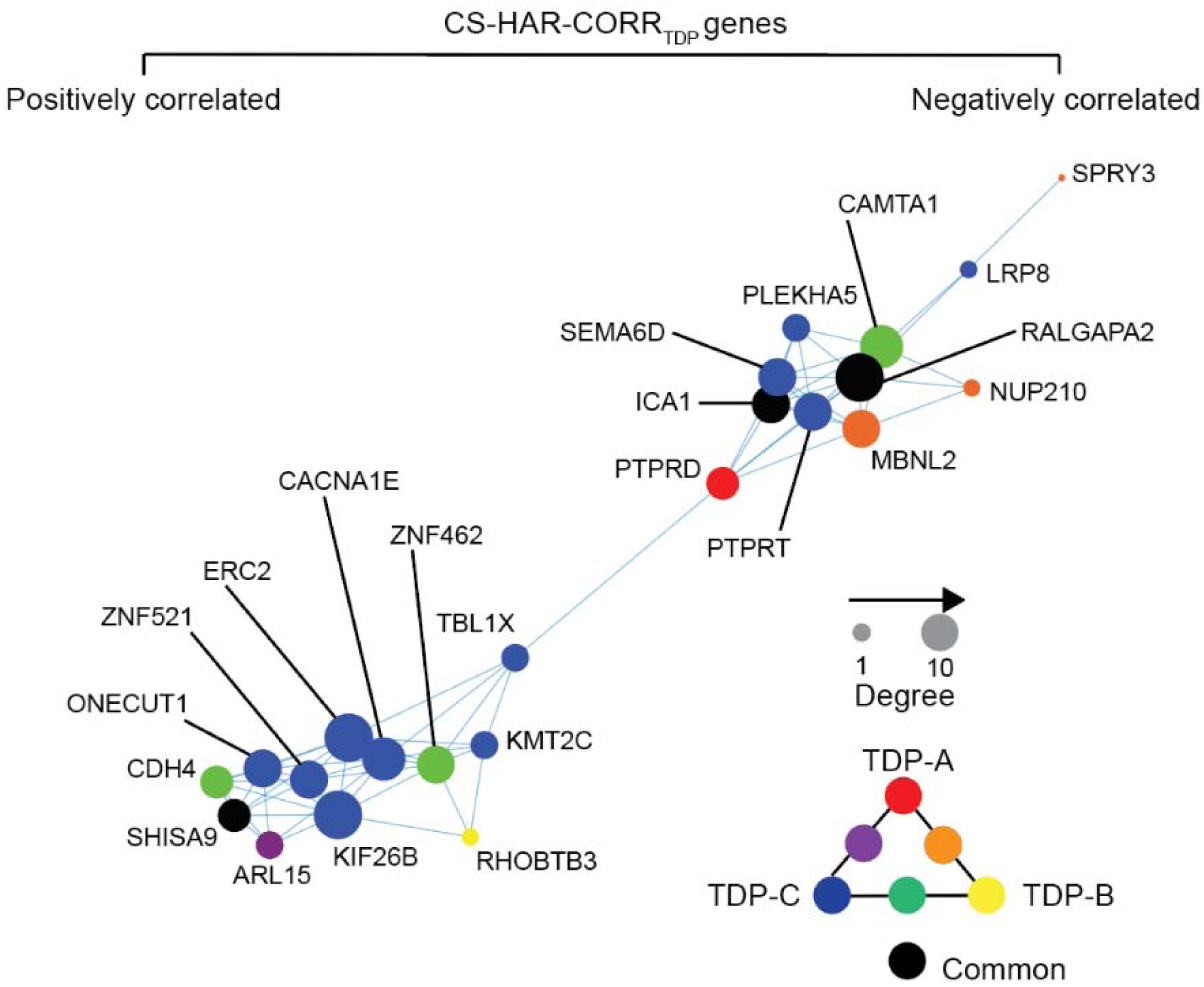
Co-expression network of CS-HAR genes associated with FTLD-TDP atrophy. The regional expression covariance of CS-HAR genes associated with FTLD-TDP atrophy was used to generate a topographical network of gene expression. The size of each circle reflects the centrality of each gene in the network, while the color of the circles reflects which FTLD-TDP subtype whose atrophy pattern correlated with the gene’s expression pattern.

### Normative expression of CS and HAR genes varies across FTLD-TDP subtypes and patient epicenters

The preceding analyses used group-level methods to identify brain-expressed HAR and CS genes associated with cortical gray matter atrophy in FTLD subtypes, but there is also prominent anatomical heterogeneity within each subtype^70^, leading us to explore a more individualized, patient-tailored approach. Neurodegenerative diseases begin within specific vulnerable and network-anchoring regions, which we have termed “epicenters” ^53,55^, before spreading to other areas along large-scale brain network connections^4,55,71^. In previous work, we introduced a method for identifying patient-specific epicenters, which enhanced network-based predictions of longitudinal atrophy progression in FTLD^53,55^. This approach uses task-free fMRI acquired in a normative sample to estimate each brain region’s connectivity to all other brain regions (***Supplementary Figure S8A***) and correlates these connectivity maps with an individual patient’s atrophy pattern to identify the region (i.e., epicenter) whose normative connectivity most resembles the atrophy pattern^53,55,56^. Based on this approach, we identified a single best-fit epicenter for each patient, focusing on FTLD-TDP given its specific relationship to cryptic splicing (**Figure 7A**). Frequency maps revealed common epicenters in TDP-A, including the pregenual and subgenual anterior cingulate cortices and the anterior insula, with the anterior cingulate being the most common epicenter. The left anterior insula was the most common epicenter in TDP-B, which showed additional epicenters in the anterior cingulate and temporal pole. In TDP-C, the left entorhinal cortex was the most common epicenter, with additional epicenters in adjacent temporal polar regions (***Supplementary Figure S8B-D***). While many epicenters were shared across subtypes (e.g., the left anterior cingulate cortex), several were associated with just one FTLD-TDP subtype, such as the left inferior frontal gyrus in TDP-A, the left angular gyrus in TDP-B, or the left fusiform gyrus with TDP-C (**Figure 7A**). Overall, despite each FTLD-TDP subtype providing 7-10 uniquely associated epicenters (**Figure 7B**), a substantial proportion of patients shared epicenters across subtypes (**Figure 7C**), suggesting that any influence of gene expression on subtype-associated regional vulnerability is not likely to be deterministic.

**Figure 7.**
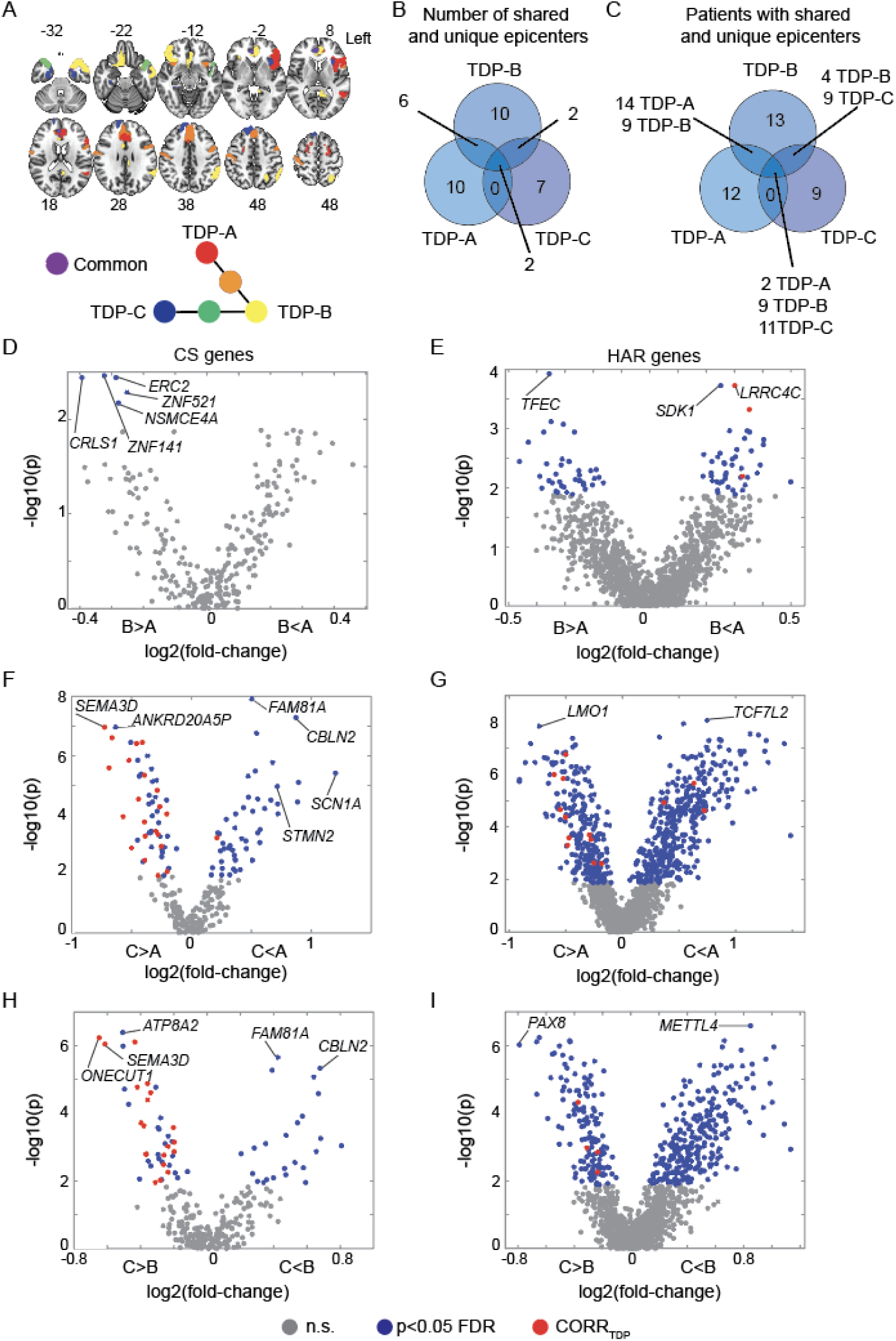
Normative expression of CS and HAR genes in FTLD-TDP disease epicenters. **(A)** Neuroanatomical distribution of FTLD-TDP subtype unique and shared epicenters. **(B)** Number of epicenters unique or shared across FTLD-TDP subtypes. **(C)** Number of patients with FTLD-TDP having unique or shared epicenters. Epicenters’ normative expression of CS genes compared between **(D)** TDP-A and TDP-B, **(E)** TDP-A and TDP-C, and **(F)** TDP-B and TDP-C. Epicenters’ normative expression of HAR genes compared between **(G)** TDP-A and TDP-B, **(H)** TDP-A and TDP-C, **(I)** TDP-B and TDP-C. Grey circles depict genes with normative expression within epicenters that do not significantly (n.s.) differ across FTLD-TDP subtypes, blue circles depict genes with normative expression within epicenters that significantly differ (FDR adjusted *p* < 0.05) across FTLD-TDP subtypes, red circles depict genes with normative expression within epicenters that significantly differ (FDR adjusted *p* < 0.05) across FTLD-TDP subtypes and also positively correlate with atrophy in the FTLD-TDP subtype where they are more expressed.

To formulate hypotheses regarding potential HAR-and CS-related genes and pathways whose expression might influence the onset of FTLD-TDP in different regions across different FTLD-TDP subtypes, we then compared the normative expression of HAR genes or TDP-43 CS genes across FTLD-TDP subtype epicenters by using Wilcoxon signed-rank tests (FDR adjusted *p < 0.05*). Resulting *p* values were plotted against the cross-fold change, reflecting higher or lower normative gene expression in epicenters of one FTLD-TDP subtype versus another (**Figure 7D-I**). These analyses revealed genes more highly expressed in regional epicenters found in TDP-B compared to TDP-A, such as CS gene *ERC2* and HAR gene *TFEC* (**Figure 7D-E**), and genes more highly expressed in epicenters for TDP-A versus TDP-B, such as HAR genes *LRRC4C* and *SDK1* (**Figure 7E**). When comparing TDP-A to TDP-C, the CS gene *STMN2* was significantly more expressed in regional epicenters found in TDP-A, while the CS gene *SEMA3D* was more expressed in regional epicenters found in TDP-C (**Figure 7F**). The HAR gene *LMO1* was significantly more expressed in epicenters of TDP-C, while the HAR gene *TCF7L2* was more expressed in epicenters of TDP-A (**Figure 7G**). When comparing TDP-B to TDP-C, the CS gene *CBLN2* and the HAR gene *METTL4* showed significantly higher normative expressions among disease epicenters derived from TDP-B, while the CS gene *ONECUT1* and the HAR gene *PAX8* were more highly expressed in epicenters of TDP-C (**Figure 7H-I**).

## Discussion

The integrative anatomical biology data presented here identified overlapping and distinct genes associated with cortical atrophy in FTLD-TDP and FTLD-tau. Both gene sets were enriched for HAR genes, providing novel comparative transcriptomic evidence that FTLD, regardless of the underlying molecular pathology, targets brain regions expressing genes that have undergone recent evolution as humans diverged from chimpanzees. Human studies leveraging chromatin interaction profiles in the fetal and adult cortex have revealed that HAR elements converge on specific cell types and laminae involved in cerebral cortical expansion^72^. These findings echo neuroimaging studies showing that the normative expression of HAR genes in the cortex is highest among brain networks that have undergone rapid evolutionary expansion in humans^45^. These recently expanded higher-order networks overlap with systems supporting language and social-emotional functions, the same systems that undergo early and progressive neurodegeneration in FTLD^70^. The normative expression of atrophy-associated HAR and non-HAR genes was either positively or negatively associated with FTLD atrophy. Genes showing a positively associated gradient are likely to be highly expressed, in the healthy brain, in regions undergoing FTLD atrophy. Dependence on these genes could contribute to FTLD regional vulnerability, especially if protein expression is diminished, for example by TDP-43 loss-of-function and associated mis-splicing. Conversely, genes displaying a negative gradient are probably expressed at lower levels, in the healthy brain, in FTLD-atrophied regions, with neurodegeneration further compromising already low levels of expression. FTLD atrophy-associated genes were characterized by common and unique associations with neuromodulatory and homeostatic transmitter systems affected in FTLD^73^. Dopaminergic, norepinephrinergic, and serotonergic imbalances are common in FTD, while the acetylcholine system remains relatively intact^59^. Common antidepressants, such as selective serotonin reuptake inhibitors, have been shown to improve the behavioral symptoms of FTD ^59^, reinforcing the notion of neuromodulatory imbalances in FTLD. Our analyses further revealed that FTLD-TDP atrophy-correlated genes are slightly enriched for genes that are targets of TDP-43 cryptic splicing repression, significantly more so than FTLD-tau atrophy-correlated genes. This observation, though somewhat tentative, suggests a potential FTLD-TDP-specific mechanism whereby incipient TDP-43 loss-of-function focuses vulnerability on specific brain networks that express high levels of TDP-43 CS genes.

Several lines of evidence derived from animal^74,75^ and cellular models^76^, as well as human neuroimaging^53,55^ and tissue studies^30^, support the notion that neuropathological changes start within vulnerable brain regions, often referred to as epicenters, before spreading via network pathways^77,78^. In this context, our study not only shows that the normative expression of HAR and CS genes resembles atrophy patterns in FTLD-TDP but also that the normative expression of these genes within putative epicenters differs among FTLD-TDP subtypes. Our findings raise the possibility that the regional expression of specific HAR and TDP-43 CS genes may predispose local circuits to cultivate specific FTLD-TDP subtypes, each characterized by a distinct TDP-43 aggregation pattern^79^. After accumulating within vulnerable epicenters, TDP-43 aggregates could then spread to other brain regions via functional and structural connectivity pathways. Histopathological studies suggest that neurodegenerative diseases may start within susceptible cell-types harbored within vulnerable brain regions^4,14,80,81^. For example, von Economo neurons and fork cells have been shown to be selectively targeted by TDP-43 depletion and inclusions in bvFTD due to FTLD-TDP^82^. These large, elongated neuron-types are almost exclusively found within the anterior cingulate and frontoinsula of large-brained, highly social mammals^83^ – including humans, greater apes, elephants, and cetaceans – raising the possibility that these neurons contribute to circuits linked to the emergence of complex social behaviors^83^. Further, a recent neuroimaging genomics study revealed that atrophied regions in FTD genetic mutation carriers express astrocyte and endothelial cell-related genes ^47^. Future histopathological studies could shed light on how expression levels of HAR and CS genes within vulnerable neuronal, such as von Economo neurons, interact with glial cells and contribute to the emergence of distinct FTLD-TDP subtypes.

Our findings should be interpreted in light of several methodological considerations. First, our study utilized normative brain transcriptomic data derived from the AHBA, which is based on bulk tissue microarray data from the brains of six donors, and both hemispheres were studied in only two of six. Future studies leveraging transcriptomic data derived from larger normative samples are needed to confirm the genetic correlates of regional gray matter atrophy in FTLD. Further, our study focused on cortical gene expression, omitting subcortical areas despite the relevance of the striatum, thalamus, and other subcortical areas in FTLD^84^. Gene expression levels from bulk tissue microarray dramatically differ depending on whether these are extracted from cortical or subcortical areas, with most studies, including ours, analyzing either one set of structures or the other^42,45^. Second, the overlap we observed between HAR and CS genes, though interesting, appears to be driven primarily by gene length. This makes sense given that longer genes may be implicated in multiple processes including brain development and neurodegenerative diseases^85,86^. Further studies are needed to assess whether altered sequences within HAR genes mechanistically interact with regions regulated or bound by TDP-43. Finally, our study focused on CS genes identified in FTLD-TDP human *postmortem* tissue data^24^ or iPSC-derived neuronal cell lines^25,26^, omitting those identified in HeLa cell lines^87^. Although this choice was intended to enhance biological confidence regarding the neural relevance of these genes, future studies may expand the TDP-43 “cryptic spliceosome” to explore a more exhaustive gene set.

## Data availability

In house Python 2.7.16 (https://www.python.org/), R 4.1.2 (https://www.r-project.org/), and MATLAB R2021a (https://www.mathworks.com/products/matlab.html) scripts were used for the analyses. W-score maps averaged for each FTLD subtype are publicly available on NeuroVault^88^. The gene lists used to generate the findings are available as ***Supplementary Data***. Further data that support the findings of this study are available on request from the corresponding author. Correspondence should be directed to Lorenzo Pasquini or William W. Seeley.

## Acknowledgements

We thank the participants and their families for their invaluable contributions to neurodegeneration research. We thank Sean Whalen for useful feedback on data analysis.

## Funding

This work was supported by NIH grants K99-AG065457 and R00-AG065457 to LP; P30AG062422, P01AG019724, U01AG057195, and U19AG063911 (WWS), as well as the Rainwater Charitable Foundation and the Bluefield Project to Cure FTD (WWS); GBHI ALZ UK-21-720973 and AACSF-21-850193 as well as JR20/0018 and PI21/00791 to IIG; 2021-A-023-FEL to AF; Gladstone Institutes and R01MH123178 to KSP.

## Competing interests

The authors declare no competing interests.

## Notes

### Competing Interest Statement

The authors have declared no competing interest.

### Author Declarations

In accordance with the declaration of Helsinki, patients or their surrogates provided written informed consent prior to participation, including consent for brain donation. The UCSF Committee on Human Research approved the study.

