## Supplement for "FTLD targets brain regions expressing recently evolved genes"

**Supplementary Materials**

- Supplementary Figures: 8
- Supplementary Tables: 1
- Supplementary Data: 1

**Supplementary tables, figures, and legends:**

| **Gene list** | **Number of genes** | **Source** |
| --- | --- | --- |
| Brain-expressed genes | 15,656 | Allen Human Brain Atlas |
| HAR genes | 2,164 | Doan et al., *Neuron* 2019 |
| CS genes 1: FTLD-TDP *postmortem* tissue | 66 | Ma et al., *Nature* 2022 |
| CS genes 2: iPSC derived neurons study I | 107 | Brown et al., *Nature* 2022 |
| CS genes 3: iPSC derived neurons study II | 233 | Sahba et al., *bioRxiv* 2023 |
| FTLD-TDP atrophy-correlated genes (merged across TDP-A, B, and C) | 8,276 | Structural MRI of 92 patients with autopsy confirmed FTLD-TDP |
| FTLD-tau atrophy-correlated genes (merged across Pick’s and CBD) | 5,580 | Structural MRI of 72 patients with autopsy confirmed FTLD-tau |

**Supplementary Table S1.** Raw numbers of gene lists involved in the analyses and their sources.

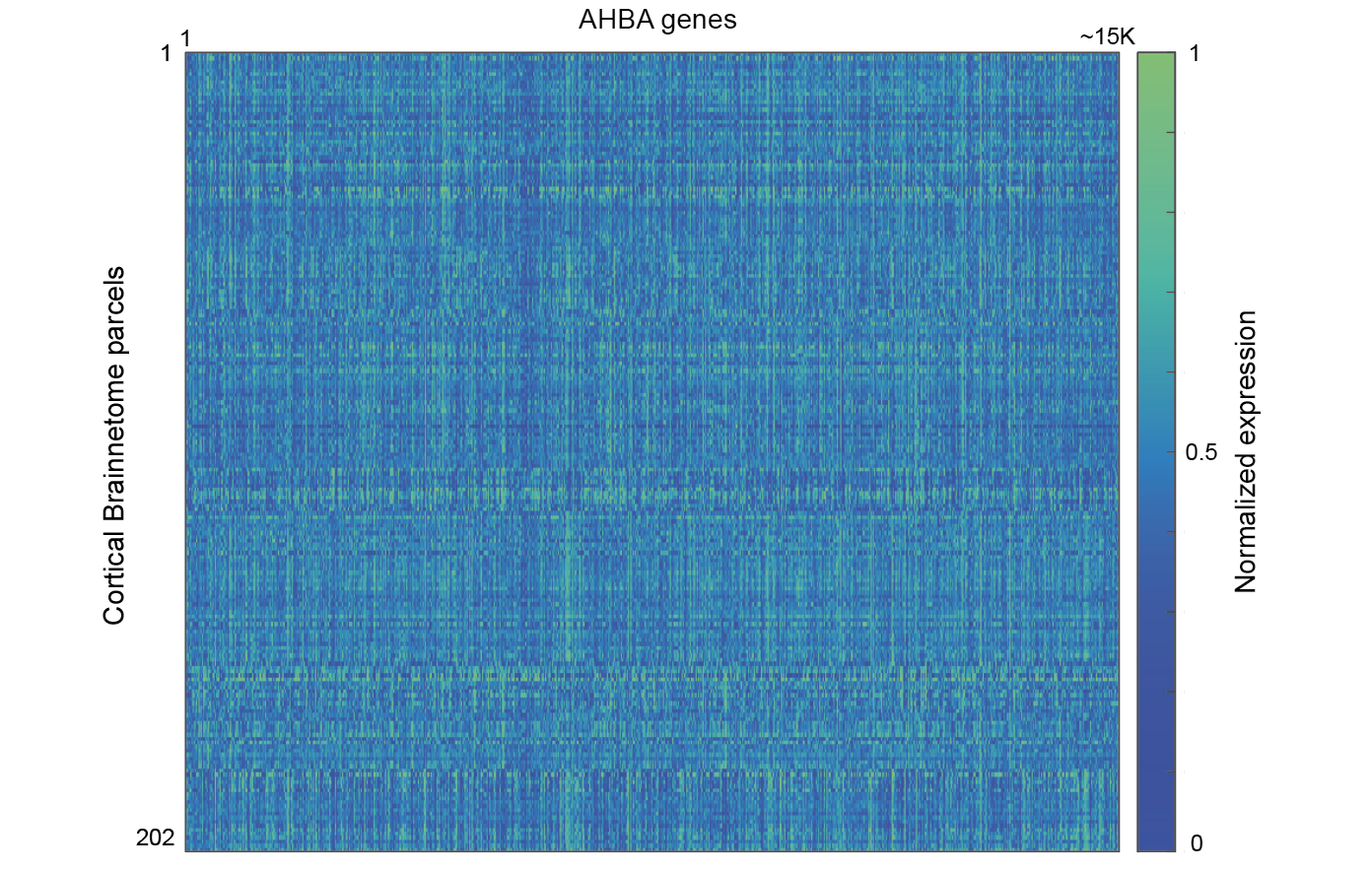

**Supplementary Figure S1. Normative gene expression across cortical parcels of the Brainnetome atlas.** **(A)** Heat matrix reflecting the normative expression of AHBA-derived brain genes within cortical parcels of the Brainnetome atlas. Color bar reflects normalized expression values (0-1).

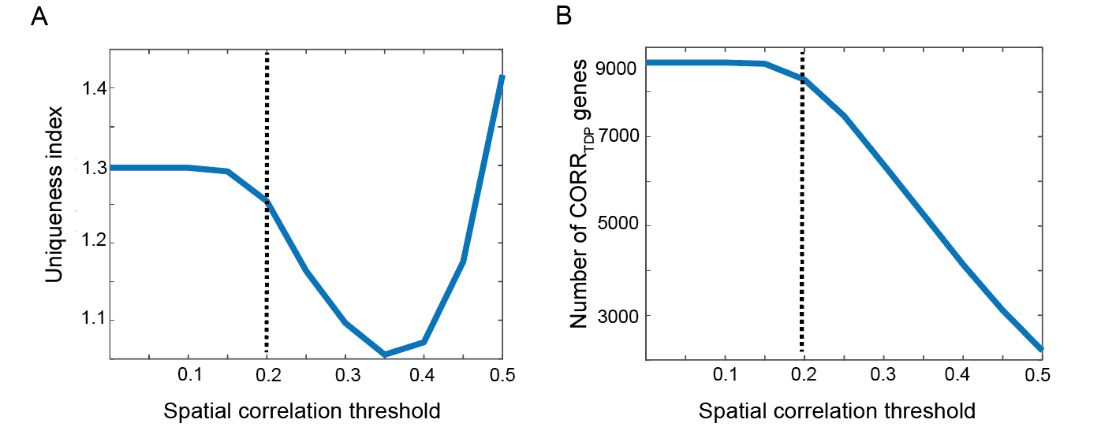

**Supplementary Figure S2. Spatial correlation threshold maximizes uniqueness and gene list length.** To remove spurious correlations from further analyses, we derived a uniqueness index reflecting the ratio between genes uniquely correlating with single FTLD-TDP subtypes and genes correlating with two or more FTLD-TDP subtypes. The optimal correlation threshold was chosen to **(A)** maximize the uniqueness index and **(B)** the number of genes correlating with FTLD-TDP subtypes.

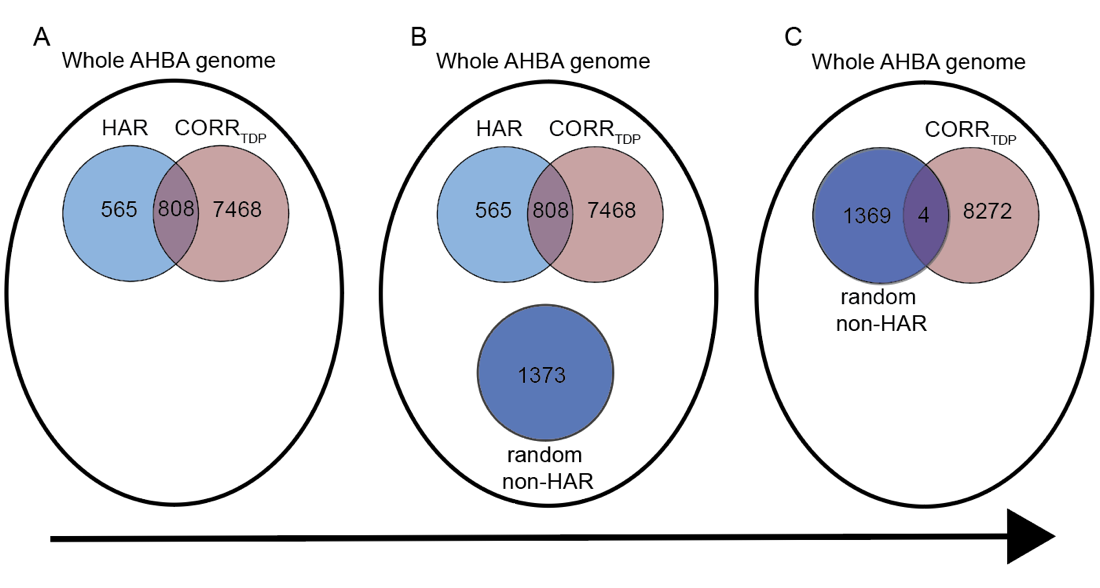

**Supplementary Figure S3. Bootstrap-based test used to assess whether the overlap between gene lists is higher than expected by chance.** **(A)** The overlap between 1,373 HAR and 8,272 FTLD-TDP atrophy correlated genes is assessed, yielding 808 common genes across both gene lists. **(B)** Across all genes expressed in the brain derived from the AHBA, 1,373 genes not overlapping with the HAR gene list are randomly selected. **(C)** the overlap between these randomly selected 1,373 genes (random non-HAR genes) and the 8,272 FTLD-TDP atrophy correlated genes is assessed. The procedure depicted in **(B)** and **(C)** is randomly repeated 5,000 times, resulting in a distribution on overlaps between randomly selected HAR genes with FTLD-TDP atrophy correlated genes, that can be compared to the observed overlap between HAR and FTLD-TDP atrophy correlated gene lists.

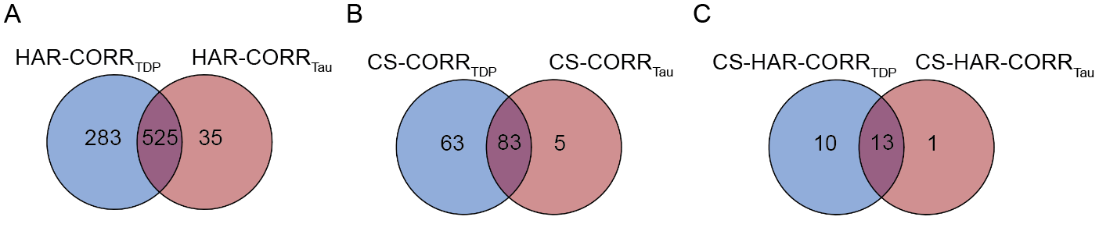

**Supplementary Figure S4. Genes correlating with atrophy in FTLD-TDP overlap with genes correlating with atrophy in FTLD-tau.** Overlaps between **(A)** HAR genes correlating with atrophy in FTLD-TDP and with atrophy in FTLD-tau; **(B)** CS genes correlating with atrophy in FTLD-TDP and with atrophy in FTLD-tau; and **(C)** CS-HAR genes correlating with atrophy in FTLD-TDP and with atrophy in FTLD-tau. Genes correlating with FTLD-TDP displayed higher levels of overlap with CS, HAR, or CS-HAR genes than genes correlating with FTLD-tau.

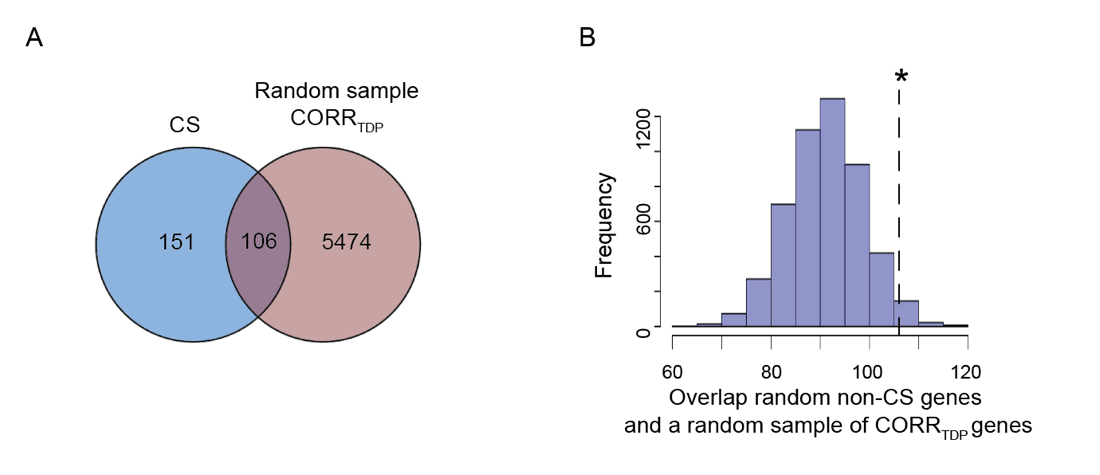

**Supplementary Figure S5. Overlap between CS genes and a random subsample of genes correlating with atrophy in FTLD-TDP. (A)** 106 genes were shared between CS genes and a randomly sampled list of 5,580 genes correlating with atrophy in FTLD-TDP. **(B)** This overlap reached significance (*p* < 0.05).

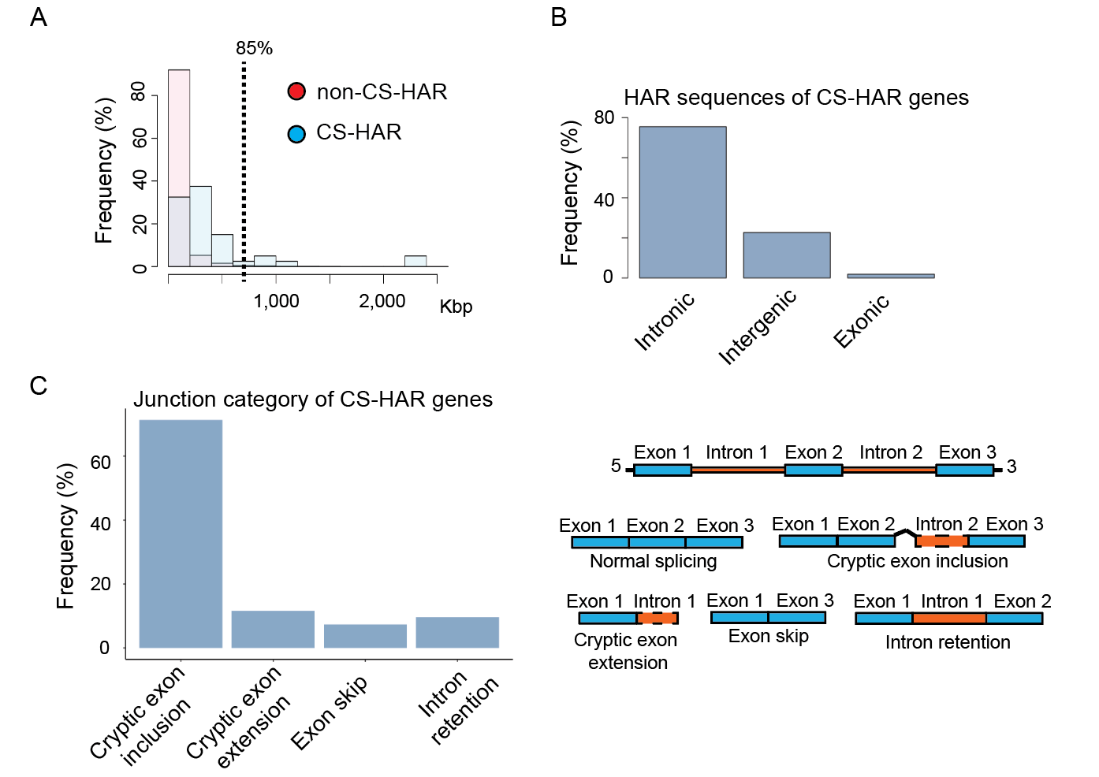

**Supplementary Figure S6. CS-HAR genes are longer than other brain expressed genes, contain mostly intronic HAR sequences, and cryptic exon cassettes are the most common mis-splicing event.** **(A)** CS-HAR genes tended to be longer than other genes in the genome, although 85% of CS-HAR genes displayed lengths comparable to those of non-CS-HAR genes. **(B)** The majority of HAR sequences of CS-HAR genes were intronic, followed by intergenic, with only a small fraction being exonic. **(C)** Junction category of miss-splicing events for genes derived from iPSC studies. The panel on the right provides a schematic overview of the distinct splicing events.

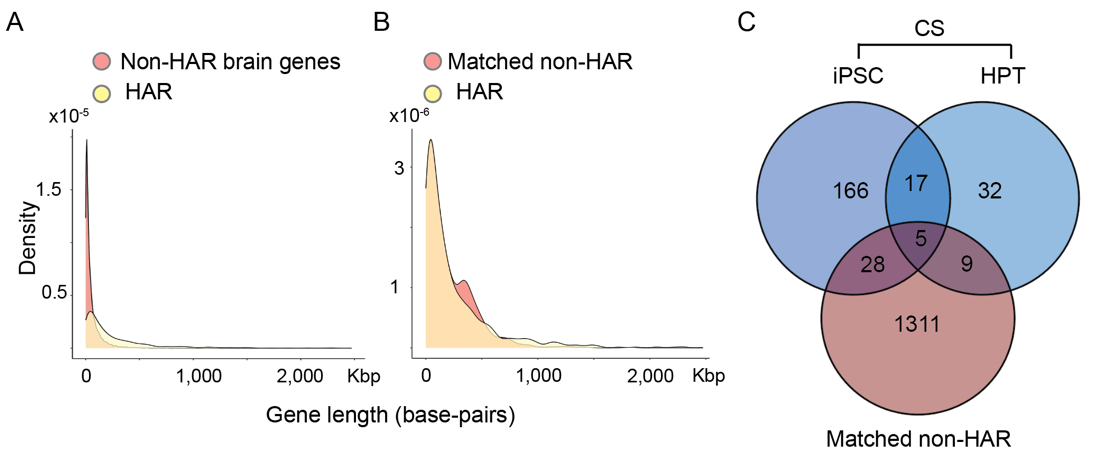

**Supplementary Figure S7. Overlap of CS genes and length-matched non-HAR genes. (A)** Length distributions of HAR brain genes and non-HAR brain genes. **(B)** Length distributions of HAR brain genes and length-matched non-HAR brain genes. **(C)** Overlap between length-matched non-HAR brain genes and CS genes.

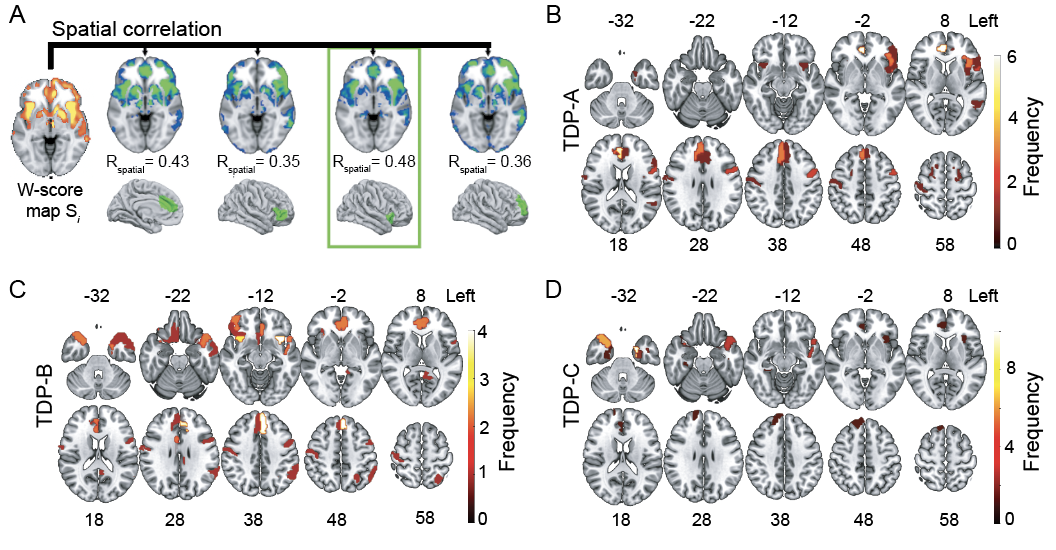

**Supplementary Figure S8. Epicenter detection and neuroanatomical distribution in FTLD-TDP subtypes. (A)** We correlated individual atrophy W-score maps with a library of functional connectivity maps derived in healthy older adults to identify epicenters in FTLD-TDP. The seed used to generate the functional connectivity map with the highest spatial similarity to the individual atrophy map was considered the epicenter for that patient. Frequency maps reflecting the spatial distribution of epicenters across FTLD-TDP subtypes. The most common disease epicenter for patients with FTLD-TDPA was in the right anterior cingulate cortex **(B)**; for FTLD-TDPB in the left anterior insula **(C)**; and for FTLD-TDPC in the left inferior temporal lobe **(D)**. Panel A adapted with permission from^53^.
